# Comparative Analysis of Reported Deaths Cases Associated with the New Coronavirus COVID-19 Pandemic in the South Caucasus Countries (Armenia, Azerbaijan, Georgia) from March 2020 to May 2022

**DOI:** 10.1101/2022.08.14.22278754

**Authors:** Avtandil G. Amiranashvili, Ketevan R. Khazaradze, Nino D. Japaridze

## Abstract

The results of a statistical analysis of daily, total by days of the week and monthly data on officially reported deaths cases from the new coronavirus COVID-19 in the countries of the South Caucasus (Armenia, Azerbaijan, Georgia) from March 12, 2020 to May 31, 2022 are presented. All data are normalized per 1 million populations (mortality rate, hereinafter, this normalization is assumed everywhere). In particular, the following results were obtained.

The daily mortality rate in Armenia averaged 3.591 (range: 0-23.622), in Azerbaijan - 1.184 (range: 0-10.969), in Georgia - 5.596 (range: 0-23.189). The total monthly mortality rate in Armenia averaged 107.9 (variability range: 1.01-415.4), in Azerbaijan - 35.6 (variability range: 0.39-122.9), in Georgia - 168.1 (variability range: 0-547.4).

A direct linear correlation was observed between the indicated countries in cases of daily and total monthly mortality.

An analysis of the intraweek course of mortality showed that in Armenia, on weekdays, the average daily mortality is 4.319, and on weekends - 3.477 (an increase compared to weekends by about 24%); in Azerbaijan, on weekdays and weekends, the average daily mortality is 1.368 and 1.421, respectively (the difference is insignificant); in Georgia on weekdays, the average daily mortality is 7.558, and on weekends - 6.855 (an increase of about 10% compared to weekends).

## 1. Introduction

More than two and a half years have passed since the outbreak of the novel coronavirus (COVID-19) in China, which was recognized as a pandemic on March 11, 2020 due to its rapid spread around the world [1]. During this period of time, despite the measures taken (including vaccination), several strains of this virus appeared (the last one is omicron and its modifications).

By the end of spring and the beginning of summer 2022, there was hope for an early end to the pandemic. However, in June-July 2022, after the last peak in January of this year, the world experienced a some increase in coronavirus cases, and in July, an increase in mortality after the last peak in February (Fig. 1, 2; data from https://data.humdata.org/dataset/total-covid-19-tests-performed-by-country). At the same time, the greatest concern is the increase in the death rate of the population.

**Fig. 1.**
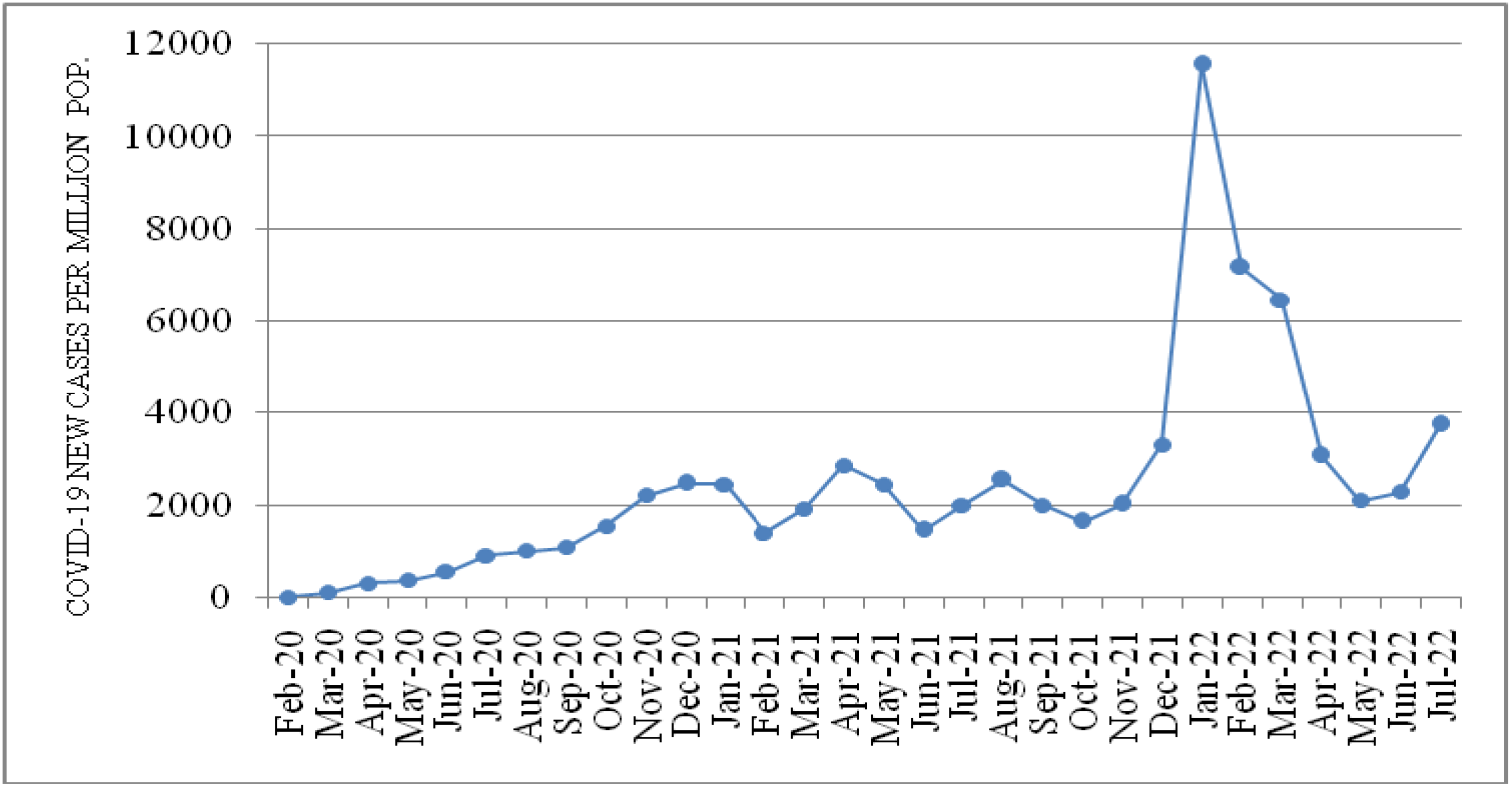
Time-series of sum monthly Covid-19 new cases per 1 million populations in World from February 2020 to July 2022.

**Fig. 2.**
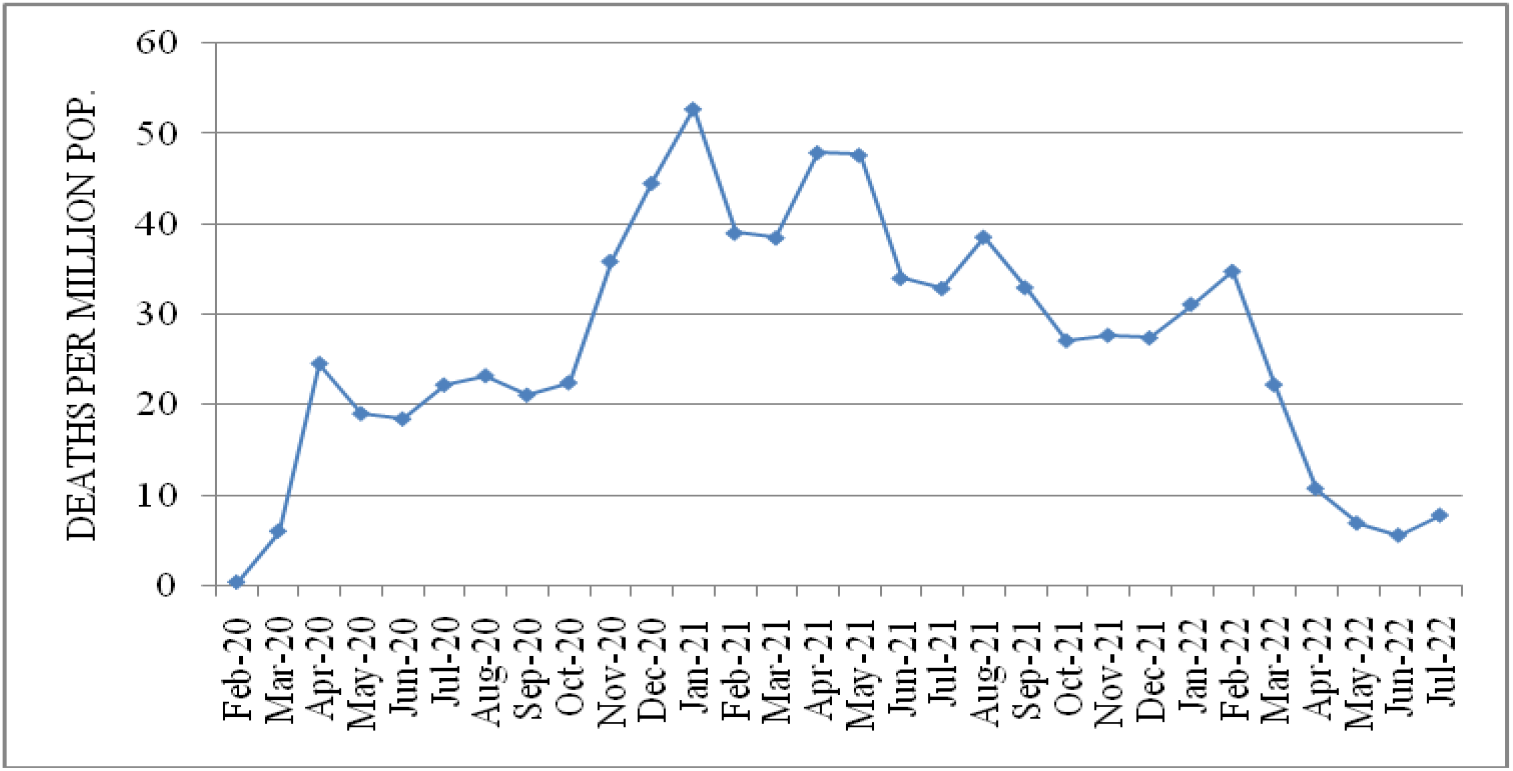
Time-series of sum monthly deaths cases from Covid-19 per 1 million populations in World from February 2020 to July 2022.

A similar situation is observed in the countries of the South Caucasus. Armenia – increase in mortality from COVID-19 in June and July (by 1.351 per 1 million populations – hereinafter, this normalization is assumed everywhere) compared to May 2022 (1.013); Azerbaijan - also an increase in mortality in June and July (0.395 and 3.064 respectively) compared to May 2022 (0.391); Georgia - increase in mortality in July (4.88) compared to June 2022 (3.524).

As before, scientists and specialists from various disciplines from all over the world continue intensive research into various aspects of the pandemic [2-17] (including in Georgia [2-15]).

This work is a continuation of the researches [8-15]. Results of a statistical analysis of the daily data associated with New Coronavirus COVID-19 confirmed deaths of the population of South Caucasus countries in the period from March 2020 to May 2022 are presented below.

It should be noted that at present, all covid-restrictions have been canceled in Georgia. Since June 2022, official data on the epidemiological situation has been published once a week, and not every day, as before. Therefore, this paper analyzes daily data on mortality from COVID-19 in the countries of the South Caucasus until May 31, 2022.

## 2. Study areas, material and methods

The study area: Armenia (ARM), Azerbaijan (AZE), Georgia (GEO). Data of John Hopkins COVID-19 Time Series Historical Data (with US State and County data) [https://www.soothsawyer.com/john-hopkins-time-series-data-with-us-state-and-county-city-detail-historical/; https://data.humdata.org/dataset/total-covid-19-tests-performed-by-country] and https://stopcov.ge about daily values of deaths coronavirus-related cases from March 12, 2020 to May 31, 2022 are used. In the proposed work the analysis of data is carried out with the use of the standard statistical analysis methods [18, 19].

The following designations will be used below: Mean – average values; Min – minimal values; Max - maximal values; Range – Max-Min; St Dev - standard deviation; CV = 100·St Dev/Mean – coefficient of variation, %; Range/ Mean, % - relative range; D – daily deaths cases per million population; Dm - sum monthly deaths cases per million population; Dw - total by days of the week deaths cases per million population; two mean values were compared using Student’s t-test with a significance level of at least 0.15; R – coefficient of linear correlation.

## 3. Results and Discussion

The results in the Fig. 3-5 and Table 1-3 are presented.

**Fig. 3.**
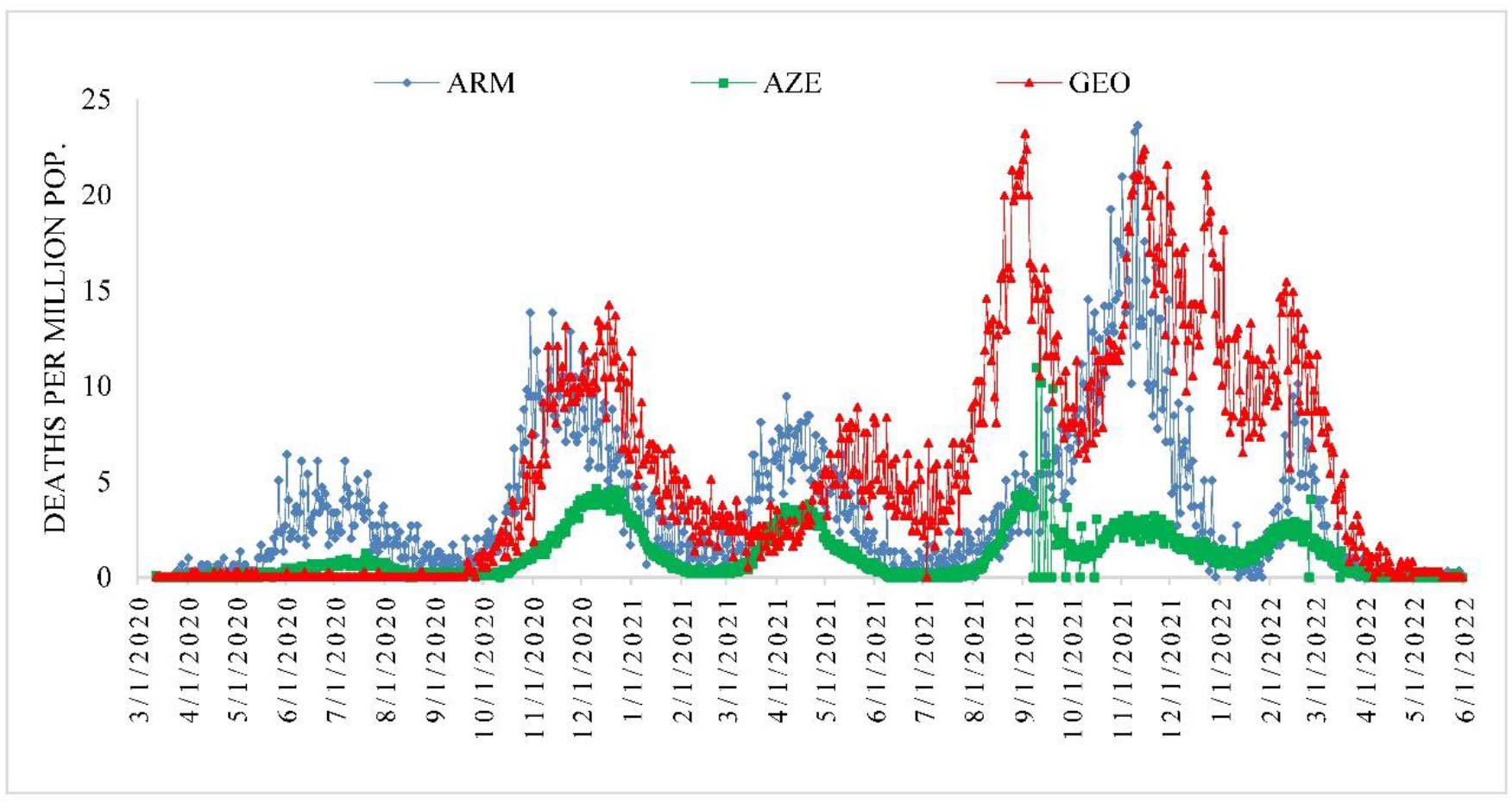
Time-series of daily deaths cases from Covid-19 per 1 million populations in Armenia, Azerbaijan and Georgia from March 12, 2020 to May 31, 2022.

**Fig. 4.**
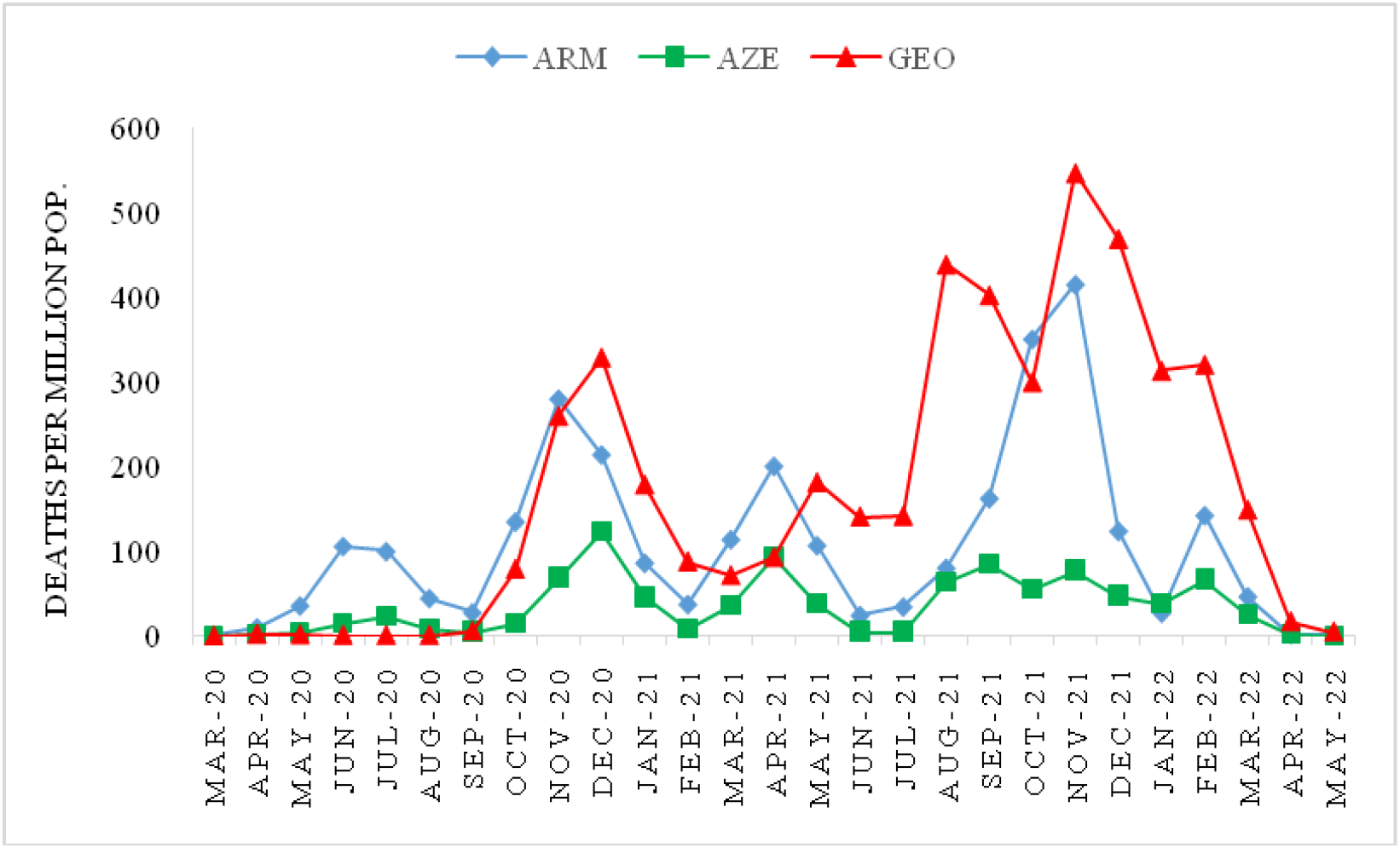
Time-series of sum monthly deaths cases from Covid-19 per 1 million populations in Armenia, Azerbaijan and Georgia from March 2020 to May 2022.

**Fig. 5.**
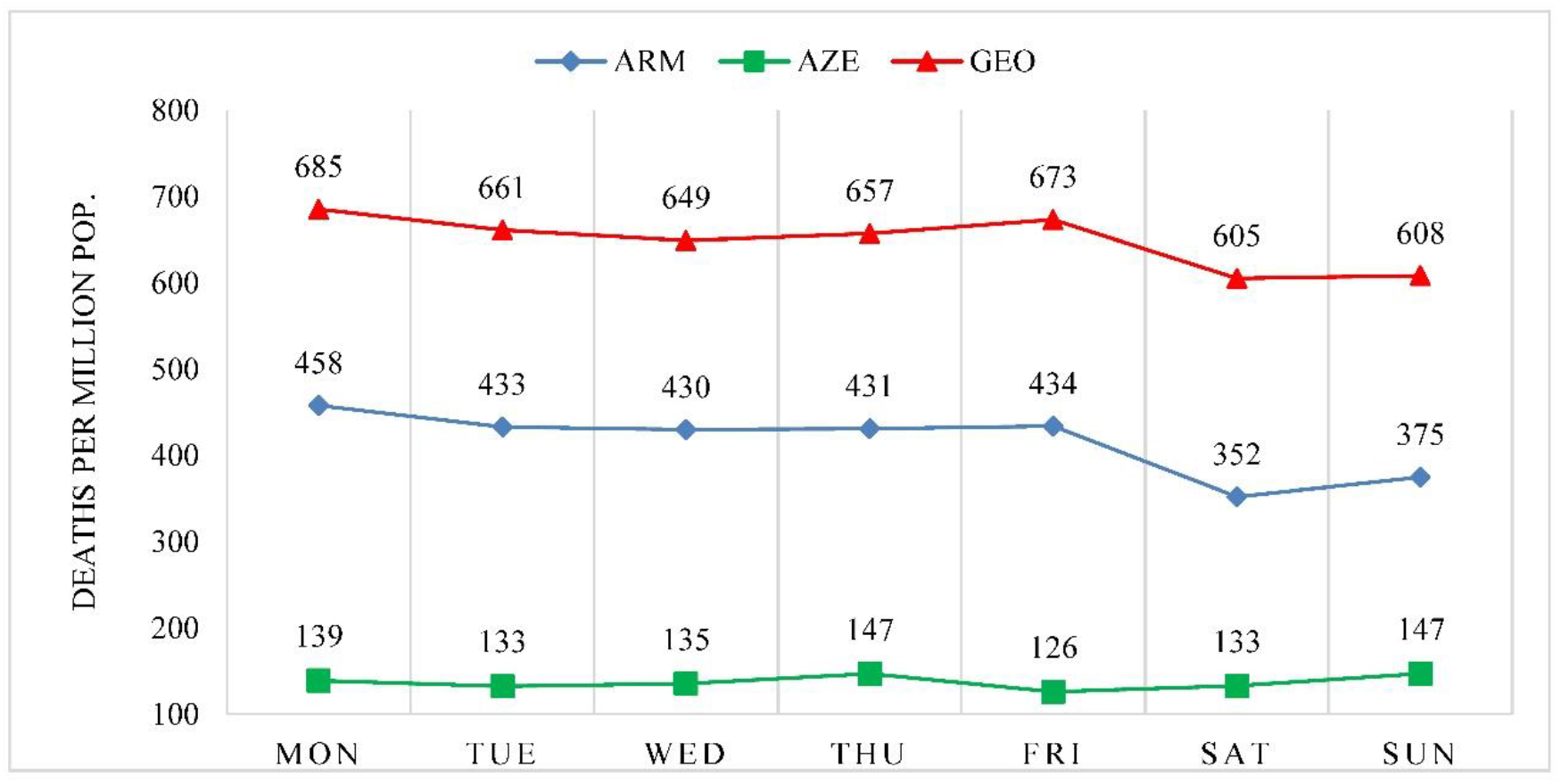
Weekly distribution of sum deaths cases from Covid-19 per 1 million populations in Armenia, Azerbaijan and Georgia from March 2020 to May 2022.

**Table 1.** The statistical characteristics of daily deaths cases from Covid-19 per 1 million populations in Armenia, Azerbaijan and Georgia from March 12, 2020 to May 31, 2022.

| Variable | ARM | AZE | GEO |
| --- | --- | --- | --- |
| Mean | 3.591 | 1.184 | 5.596 |
| Min | 0 | 0 | 0 |
| Max | 23.622 | 10.969 | 23.189 |
| St Dev | 3.879 | 1.329 | 5.795 |
| Cv, % | 108.0 | 112.3 | 103.5 |
| Range/Mean, % | 658 | 927 | 414 |
| Correlation matrix |  |  |  |
| ARM | 1 | 0.58 | 0.55 |
| AZE | 0.58 | 1 | 0.62 |
| GEO | 0.55 | 0.62 | 1 |

**Table 2.** The statistical characteristics of sum monthly deaths cases from Covid-19 per 1 million populations in Armenia, Azerbaijan and Georgia from March 2020 to May 2022.

| Variable | ARM | AZE | GEO |
| --- | --- | --- | --- |
| Mean | 107.9 | 35.6 | 168.1 |
| Min | 1.01 | 0.39 | 0.0 |
| Max | 415.4 | 122.9 | 547.4 |
| Range | 414.4 | 122.5 | 547.4 |
| St Dev | 106.8 | 33.9 | 168.1 |
| Cv, % | 99.0 | 95.3 | 100 |
| Range/Mean, % | 384 | 344 | 326 |
| Correlation matrix |  |  |  |
| ARM | 1 | 0.73 | 0.62 |
| AZE | 0.73 | 1 | 0.73 |
| GEO | 0.62 | 0.73 | 1 |

**Table 3.** The statistical characteristics of sum weekly deaths cases from Covid-19 per 1 million populations in Armenia, Azerbaijan and Georgia from March 2020 to May 2022.

| Variable | ARM | AZE | GEO |
| --- | --- | --- | --- |
| Mean | 416 | 137 | 648 |
| Min | 352 | 126 | 605 |
| Max | 458 | 147 | 685 |
| Range | 106 | 21 | 80 |
| St Dev | 37.8 | 7.8 | 30.8 |
| Cv, % | 9.1 | 5.7 | 4.7 |
| Range/Mean, % | 25.4 | 15.4 | 12.3 |
| Correlation matrix |  |  |  |
| ARM | 1 | -0.12 | 0.97 |
| AZE | -0.12 | 1 | -0.27 |
| GEO | 0.97 | -0.27 | 1 |

Fig. 3 shows the time course of the daily mortality rate D in Armenia, Azerbaijan and Georgia in the period from March 12, 2020 to May 31, 2022. As follows from this figure, the time course of this indicator for all the countries studied was undulating. At the same time, in Armenia and Georgia there were five pronounced peaks in the mortality rate, in Azerbaijan - six.

The peaks of the daily mortality rate are sometimes close in time, sometimes significantly shifted relative to each other. So, for example, the peak of the first wave of the daily mortality rate in Armenia (Jun 1, 2020, D=6.416) was 50 days ahead of the peak of the first wave of this indicator in Azerbaijan (Jul 20, 2020, D=1.291) and more than 5.5 months in Georgia (Dec 18, 2020, D=14.237). The peaks of the second wave of the daily mortality rate in Armenia (Oct 30 and Nov 13, 2020, D=13.844) occurred 41 and 27 days earlier than the peak of the second wave of deaths in Azerbaijan (Dec 10, 2020, D=4.669) and more than 5 months earlier than in Georgia (May 21, 2021, D=8.898).

The peak of the third wave of the death rate in Armenia (Apr 7, 2021, D=9.449) was close in time to the third peak of the death rate in Azerbaijan (Apr 21, 2021, D=3.854) and almost 5 months ahead of the peak of the third wave of the daily death rate in Georgia (Sep 2, 2021, D=23.189). The fourth highest peak in the death rate in Armenia (Nov 11, 2021, D=23.622) was about two months behind the peak in Azerbaijan (Sep 9, 2021, D=10.969) and was close in time to the peak in Georgia (Nov 15, 2021, D=22.380). The fifth peak of mortality in Azerbaijan was observed on Nov 5 and Nov 21, 2021 (D=3.261). The fifth peak of mortality in Armenia (Feb 18, 2022, D=10.130), the sixth peak of mortality in Azerbaijan (Feb 26, 2022, D=4.108) and the fifth peak of mortality in Georgia (Feb 11, 2022, D=15.453) were observed in relatively close time interval.

Table 1 presents the statistical characteristics of the daily mortality rate in the countries studied.

As follows from this table, the average daily mortality rate in Armenia was 3.591 (range: 0-23.622), in Azerbaijan - 1.184 (lowest, range: 0-10.969), in Georgia - 5.596 (highest, range: 0-23.189). On average, the daily mortality rate in Georgia was 1.56 times higher than in Armenia and 4.73 times higher than in Azerbaijan. The time series of the daily mortality rate in all countries is subject to strong variations. At the same time, the largest variations of this indicator were observed in Azerbaijan (Cv=112.3%, Range/Mean=927%), the smallest - in Georgia (Cv=103.5%, Range/Mean=414%).

A direct linear correlation (moderate correlation) was observed between these countries in terms of daily deaths, which varied from 0.55 (pair: Armenia-Georgia) to 0.62 (pair: Azerbaijan-Georgia).

Fig. 4 shows the time course of the total monthly mortality rate in Armenia, Azerbaijan and Georgia in the period from March 2020 to May 2022. As in the previous case, the time course of this indicator for all the countries studied was undulating with five peaks of this mortality rate in Armenia and Georgia and six - in Azerbaijan.

As for the daily mortality rate, the peaks of the total monthly mortality rate in some cases coincide in time, and in some cases are shifted relative to each other. The first peak of the total monthly mortality rate in Armenia (Jun 2020, Dm=106.0) is one month ahead of the first peak in Azerbaijan (Jul 2020, Dm=23.2) and 5 months ahead of Georgia (Dec 2020, Dm=329.1). The second peak of the total monthly mortality rate in Armenia (Nov 2020, Dm=280.3) was observed a month earlier than the peak of this indicator in Azerbaijan (Dec 2020, Dm=122.9) and 5 months earlier than in Georgia (May 2021, Dm=181.7).

The peaks of the third wave of the total monthly mortality rate in Armenia and Azerbaijan coincided and were observed in April 2021 (Dm=200.8 and Dm=93.4 respectively) and were 3 months ahead of the peak of the third wave of the indicated total monthly mortality rate in Georgia (Aug 2021, Dm=439.2) The fourth, largest peak in the mortality rate in Armenia (Nov 2021, Dm=415.4), was two months behind the peak in mortality in Azerbaijan (Sep 2021, Dm=85.6) and coincided in time with the peak in mortality in Georgia (Nov 2021, Dm=547.4). The fifth peak of total monthly mortality in Azerbaijan was observed in November 2021 (Dm=77.4). The fifth peak of total monthly mortality in Armenia and Georgia and the sixth of this indicator in Azerbaijan was observed in February 2022 (Dm=142.5, 320.7 and 66.8 respectively).

Table 2 presents the statistical characteristics of the total monthly mortality rate in the countries studied.

As follows from this table, the monthly mortality rate in Armenia averaged 107.9 (range of variation: 1.01-415.4), in Azerbaijan - 35.6 (lowest indicator, range of variability: 0.39-122.9), in Georgia - 168.1 (highest indicator, range of variability: 0.0-547.4). As in the previous case (table 1), the time series of the total monthly mortality rate in all countries are subject to strong variations. The highest value of the coefficient of variation was noted in Georgia (Cv=100%, Range/Mean=927%), the lowest - in Azerbaijan (Cv=95.3%). The highest value of the relative variation range was noted in Armenia (Range/Mean=384%), the smallest - in Georgia (Range/Mean=326%).

A direct linear correlation was observed between these countries in terms of total monthly mortality, which was 0.62 (pair: Armenia-Georgia, moderate correlation) and 0.73 (pairs: Armenia-Azerbaijan and Azerbaijan-Georgia, high correlation).

An analysis of data on indicators of total mortality in the countries of the South Caucasus by days of the week showed (Fig. 5) that the maximum mortality in Armenia and Georgia was observed on Monday (458 and 685, respectively), the minimum - on Saturday and Sunday (352 and 375, and 605 and 608, respectively). In Azerbaijan, the maximum total mortality was observed on Thursday and Sunday (147 each), the minimum on Friday (126).

In general, in Armenia, on weekdays, the average daily mortality is 4.319, and on weekends - 3.477 (an increase compared to weekends by about 24%); in Azerbaijan, on weekdays and weekends, the average daily mortality is 1.368 and 1.421, respectively (the difference is insignificant); in Georgia on weekdays, the average daily mortality is 7.558, and on weekends - 6.855 (an increase of about 10% compared to weekends). We also note that in Azerbaijan on Thursday and Sunday (the highest mortality compared to the rest of the days of the week, Fig. 5) the average daily mortality is 1.455, and on the other days of the week - 1.354 (the difference is also insignificant, as for weekdays and weekends).

It should be noted that data on the intraweek dynamics of mortality from coronavirus for some countries completely contradict our data for Armenia and Georgia. For example, in [16] the average number of deaths on weekends with the average number of the preceding 5 days for 10 countries with the highest number of COVID-19 cases was compared (the United States, United Kingdom, France, Germany, Italy, Spain, Russia, India, Brazil and Canada). On average over the 2-year study period, more people died on a weekend day (average daily number of global COVID-19 deaths - n = 8.532) than a weekday (n = 8.083).

Among the 10 countries with the highest case counts, all but Germany reported higher death averages on the weekend when compared with weekdays. The highest absolute increase in weekend deaths occurred in the U.S. (22% increase), Brazil (29% increase) and the U.K. (11% increase).

To identify the causes of these discrepancies, special studies will be required.

Compared with the values of D and Dm, the variations in total mortality by day of the week are much lower. The largest variations in the Dw parameter were noted in Armenia (Cv=9.1%, Range/Mean=25.4%), the smallest - in Georgia (Cv=4.7%, Range/Mean=12.3%). In Azerbaijan, the values of the coefficient of variation and the relative variation range were 5.7% and 15.4%, respectively (Table 3).

It also follows from Table 3 that according to the Dw values, a very high direct linear correlation (R=0.97) was observed between Armenia and Georgia. For Armenia-Azerbaijan and Azerbaijan-Georgia pairs, R = -0.12 and -0.27 (negligible correlation).

In conclusion, we note that the real data on mortality from COVID-19 may be much higher than officially confirmed. In [17], it is indicated that mortality varies by time and location, and its measurement is affected by well known biases that have been exacerbated during the COVID-19 pandemic. Authors of this paper to estimated excess mortality from the COVID-19 pandemic in 191 countries and territories, and 252 subnational units for selected countries, from January 1, 2020, to December 31, 2021.

By December 31, 2021, global reported deaths due to COVID-19 reached 5.94 million, but the estimated number of excess deaths was nearly 3.07-times (95% UI 2. 88–3.30) greater, reaching 18.2 million (17.1–19.6). The global all-age rate of excess mortality due to the COVID-19 pandemic was 120.3 deaths (113.1–129.3) per 100 000 of the population.

Wherein, ratio between excess mortality rate and reported COVID-19 mortality rate in Armenia is 2.59 (from 2.46 to 2.75), in Azerbaijan – 6.40 (from 6.15 to 6.66) and in Georgia – 1.15 (from 1.02 to 1.30) [17].

In the near future, we are going to conduct a similar assessment of excess mortality from COVID-19 for the countries of the South Caucasus by comparing the actual data on total mortality in these countries in 2020-2021 with extrapolated data suggesting the absence of a pandemic and to compare our assessments with the results of work [17].

## Conclusion

In the future, until the full end of the pandemic, we will continue similar studies for Georgia in comparison with neighboring and other countries.

## Data Availability

https://www.soothsawyer.com/john-hopkins-time-series-data-with-us-state-and-county-city-detail-historical/; https://data.humdata.org/dataset/total-covid-19-tests-performed-by-country; https://stopcov.ge

https://www.facebook.com/Avtandil1948/

https://www.soothsawyer.com/john-hopkins-time-series-data-with-us-state-and-county-city-detail-historical/

https://data.humdata.org/dataset/total-covid-19-tests-performed-by-country

https://stopcov.ge

